# COVID-19 Vaccine effectiveness against symptomatic infection and hospitalization in Belgium, July 2021-APRIL 2022

**DOI:** 10.1101/2022.05.09.22274623

**Authors:** Toon Braeye, Joris van Loenhout, Ruben Brondeel, Veerle Stouten, Pierre Hubin, Matthieu Billuart, Jenny Chung, Mathil Vandromme, Chloé Wyndham-Thomas, Koen Blot, Lucy Catteau

## Abstract

The COVID-19 vaccination campaign in Belgium aimed to reduce disease spread and severity. We quantified the observed vaccine effectiveness (VE) against symptomatic infection (VEi) and hospitalization (VEh).

Exhaustive data on testing and vaccination was combined with a clinical hospital survey. We estimated VEi using a test negative design and VEh using a proportional hazard analysis. We controlled for prior infection, age, sex, province of residence and calendar week of sampling. Variant of concern specific VE-estimates were obtained by time since vaccination from July 2021 to April 2022.

We included 1,433,135 persons. VEi against Delta waned from an initial estimate of 81% (95%CI 80- 82) to 56% (95%CI 56-57) 100-150 days after primary-vaccination. Booster-vaccination increased initial VEi to 84% (95%CI 83-85). Against Omicron, an initial VEi of 37% (95%CI 34-40) waned to 18% (95%CI 17-20) 100-150 days after primary-vaccination. Booster-vaccination increased VEi to 52% (95%CI 51-53) and waned to 25% (95%CI 24-27) 100-150 days after vaccination. Hybrid immunity conferred by prior infection and booster-vaccination outperformed booster-vaccination only even if the infection was over one year ago, 67% (95%CI 66-68). Initial VEh for booster-vaccination decreased from 93% (95%CI 93-94) against Delta to 87% (95%CI 85-89) against Omicron. VEh for Omicron waned to 66% (95%CI 63-70) 100-150 days after booster-vaccination.

In conclusion, we report significant immune-escape by Omicron. VEh was less affected than VEi and immune-escape was attenuated by booster-vaccination. Waning further reduced VEi- and VEh- estimates. Infection-acquired immunity offered additional protection against symptomatic infection in vaccinated persons which lasted at least one year.

## 2. Introduction

Belgium started the roll-out of its COVID-19 vaccination campaign on 28 December 2020. When a third wave of high COVID-19-incidence occurred in March and April 2021 associated with the Alpha Variant Of Concern (VOC), nursing home residents and a proportion of healthcare professionals had completed the primary vaccination schedule (primary-vaccination). A fourth wave associated with the Delta-VOC started in October 2021, despite a vaccination coverage of more than 80% in the adult (≥18 years) population. Delta showed immune-escape which, in combination with waning of vaccine-induced protection, lowered vaccine effectiveness (VE) against infection [1]. The fourth wave resulted in considerable ICU and hospital occupancy. When the fourth wave subsided in December 2021, it was quickly followed by a fifth wave in January 2022 associated with the Omicron-VOC. Throughout the fourth wave, the booster campaign which started in September 2021, was sped up. Its relevance was increased even further due to the requirement of a booster dose to effectively neutralize Omicron [2]. Vaccination centers were scaled up and the interval between primary and booster-vaccination was shortened mid-December 2021 from six to four months for mRNA-vaccines. The interval between a viral-vector vaccine based priming and a booster shot remained at two months (for Ad26.COV2.S) and four months (for ChAdOx1). On 28 February 2022, 88.9% of all Belgians aged 18 years and above had completed primary-vaccination and 74.4% had received a booster dose.

Several trends characterized the ending of 2021 and start of 2022: increasing vaccination coverage, a change in dominant VOC and a large increase in the cumulative number of laboratory-confirmed infections up to 3.21 million for 11.6 million residents on 31 January 2022. The effects of vaccination are confounded within these trends. Observational [3–6] and *in-vitro* studies [7] reported that the effectiveness of vaccines decreases over time, a process typically referred to as waning, and that several VOCs have shown some degree of immune-escape. Emerging variants have also been associated with an increased risk of reinfection and this appears to be particularly true for Omicron [8,9]. Given these findings, VE cannot be directly interfered from an observed trend without adjusting for VOC, prior infections and time since vaccination. Timely and detailed estimates of VE however are essential to further develop and adjust the vaccination strategy.

In the present study, we estimated VE against symptomatic infection (VEi) and hospitalization (VEh) associated with primary and booster-vaccination. In addition, we estimated protection offered by hybrid immunity, i.e. immunity resulting from the combination of SARS-COV-2 infection and vaccination.

## 3. Methods

### 3.1. DATASET & STUDY PERIOD

This study was undertaken in the architecture of the LINK-VACC project. It allows the linkage of selected variables from multiple existing national health and social sector registries on the individual level using the Belgian social security identification number within a pseudonymized environment. Three COVID-19 registries were used for the present study: the vaccination registry, the laboratory testing registry and the clinical hospital survey. The vaccination (sex, age, place of residence, date of vaccination, vaccination brand) and laboratory testing (date of sampling, test result, presence of symptoms) registries are exhaustive. The clinical hospital survey (date and reason of admission) is based on a voluntary survey filled in by the hospitals and contains 41% (18,886/45,999) of all hospital admissions for COVID-19 in Belgium during the study period. Persons hospitalized with a laboratory SARS-CoV2-confirmed infection but for whom the reason for admission was not linked to COVID-19- symptoms were excluded.

Belgium predominantly used two viral vector vaccines (ChAdOx1 and Ad26.COV2.S) for primary-vaccination and two mRNA vaccines (BNT162b2 and mRNA-1273) for both primary and booster-vaccination. We analysed VE by completion of primary-vaccination (two doses except for Ad26.COV2.S for which one dose suffices) or booster-vaccination. Persons partially vaccinated (one-dose of a two-dose schedule) or vaccinated with a mixed primary-vaccination schedule were excluded. A dose of an mRNA-vaccine administered after primary-vaccination was considered booster-vaccination.

Since relatively few laboratory-confirmed infections were sequenced (5.4% during the study period), the time period during which the sample was taken was used as a proxy for Delta and Omicron-infections. The GISAID-database was used to define these dominant periods. If one VOC was detected in at least 80% of the sequenced samples during a period, this VOC was considered dominant. We did not differentiate between the different Omicron sublineages, as equal immunogenicity against Omicron sublineages (BA.1, BA.2) has been reported [10]. By this definition, the Delta-period started July 15 2021 and ended December 6 2021 and the Omicron-period included in this study started January 3 2022 and ended April 14 2022 (end of the study period). We included all Belgian adults (≥18 years old) with a PCR-test for SARS-CoV2-infection after symptom onset. We excluded tests that were performed for the confirmation of a positive self-test, because having a positive self-test possibly already depends on vaccination.

### 3.2. IMMUNITY STATUS: VACCINATION AND PRIOR INFECTION

Immunity status was defined by: (1) the vaccination status: unvaccinated, primary or booster-vaccination, (2) time since vaccination in 50-day blocks (from the date on which the last administered vaccine was considered effective) and (3) most recent prior infection. Prior infections were categorized by time of sampling: before 2021, first half of 2021, second half of 2021 and since 2022. A positive test within 60 days of a previous positive test were discarded as prior infection as they possibly reflected the same infection.

ChAdOx1 and mRNA-1273 vaccines were considered effective 14, BNT162b2 seven and Ad26.COV2.S 21 days after administration. Booster doses were considered effective seven days after administration.

### 3.3. TEST NEGATIVE DESIGN

In the test-negative design, immunity status was compared between symptomatic persons who tested positive for SARS-CoV2 (cases) and those who tested negative (controls). Cases and controls were matched on age group (per 5 years), sex, province of residence and calendar week of sampling.

Since information on symptoms was self-reported and we assumed ‘between person’-variation with respect to self-reported symptoms and testing behaviour, we chose not to include multiple symptomatic periods per person, but instead randomly chose one symptomatic period per person. A symptomatic period was defined by a symptom-onset-date as reported by the person during sampling. If all PCR-tests within 20 days of the onset-date were negative, this person was considered negative; otherwise (one or multiple positive PCR-tests), the person was considered positive.

Data on test results was fitted using conditional logistic regression to obtain immune status-specific adjusted odds ratios (aOR). Unvaccinated persons without a prior infection were set as reference category. VEi was estimated as (1-aOR)*100.

### 3.4. PROPORTIONAL HAZARD ANALYSIS

To estimate additional VE against hospitalization given symptomatic infection (VEh|i), we combined person-level data on the immunity status of persons with a symptomatic infection with a clinical hospital survey. Persons with a symptomatic infection had a follow-up of four weeks to see if hospitalization occurred. Persons were censored from follow-up if they died or changed immunity status (e.g. received booster-vaccination).

We estimated the hazard ratio (HR) for hospital intake for COVID-19 by immunity status while adjusting for age group (per 5 years), sex and province of residence using Cox-regression. VEh|i is estimated as (1-HR)*100. Persons with a confirmed prior infection were excluded from the Cox-regression because of small numbers. Only descriptive statistics were reported for hospitalization of convalescents.

VEh was obtained by combining VEi and VEh|i.

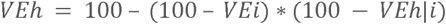

### 3.5. MODEL FIT & STRATIFICATION

Models were fitted on datasets stratified by VOC-period. When we report VE-estimates for an age group (e.g. over 65 years of age) the dataset used for coefficient estimation was limited to persons of that age group, but within the model we adjusted for the 5-year age groups previously mentioned. CI refers to 95% confidence intervals. R 4.1.2 and the R-package ‘survival’ were used for the model fit. To improve the readability of the graph and to avoid over-interpretation, we did not plot estimates for which the uncertainty was large (CI > 50%).

## 4. Results

### 4.1. NUMBER OF OBSERVATIONS INCLUDED IN THE ANALYSIS

The number of PCR-tests performed during the study period was 14,397,138 (of which 2,780,968 were positive, 19%) linked to 6,703,888 persons. We excluded 11,624,075 PCR-tests because they were linked to asymptomatic persons (3,809,815) or because no information on symptoms was available (7,814,260). The remaining 2,773,063 PCR tests were linked to 2,646,797 unique symptomatic periods from 2,123,900 persons.

We excluded tests from minors (<18 years, N=585,018). Among adults, tests were further excluded because of (multiple reasons might apply) incomplete primary-vaccination (N=44,751), non-standard/mixed primary-vaccination (N=2,555), PCR-test after a positive self-test (N=98,203) or because the sample was collected in between the Delta and Omicron-period (N=176,072).

A total of 1,433,135 persons were included in the VEi-analysis (Table 1). For the analysis of VEh|i, 662,220 symptomatic, infected persons were included (table 2).

**Table 1:**
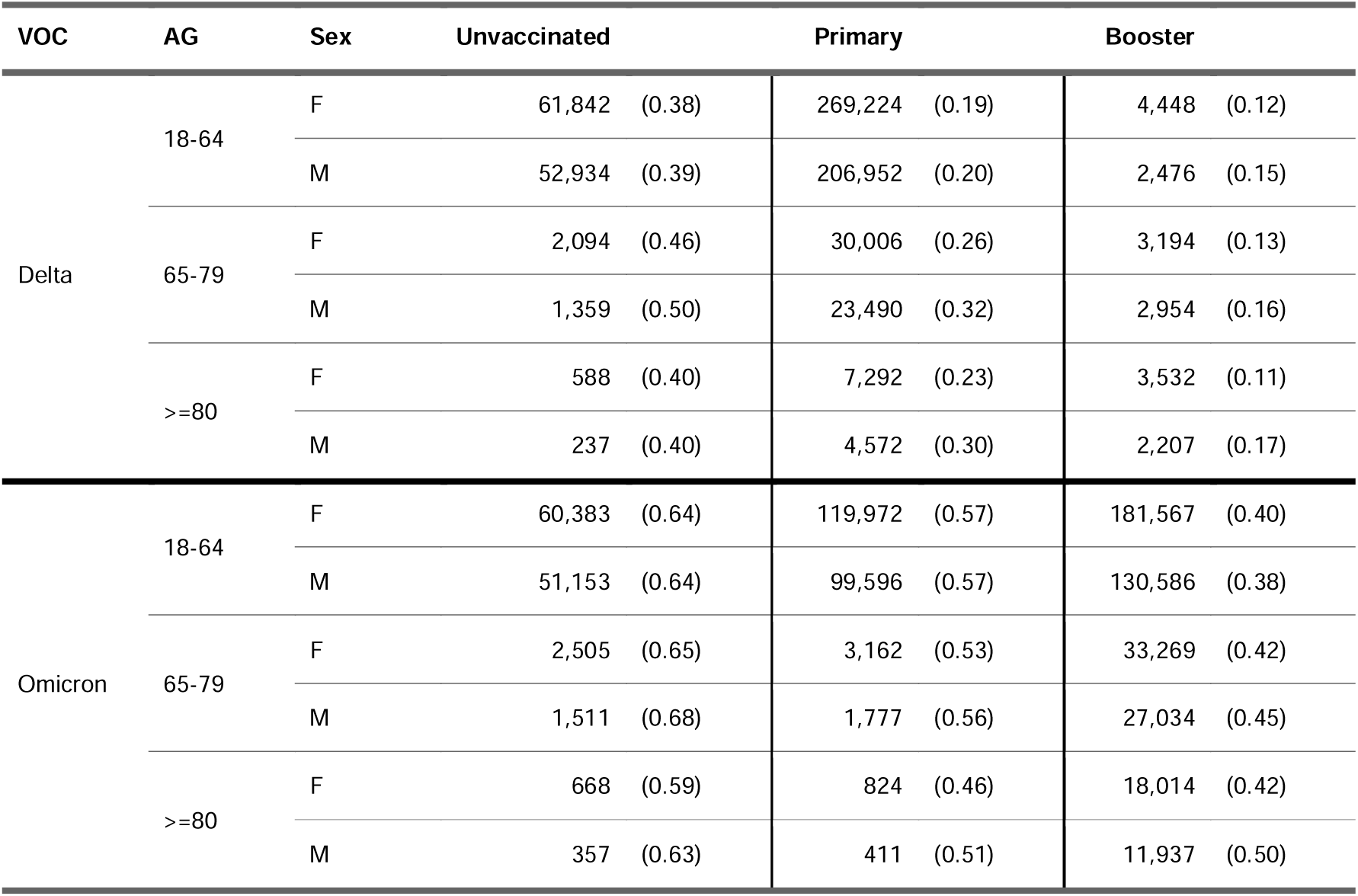
Number of persons included in the test-negative study by Variant of Concern (VOC), Age Group (AG), sex and vaccination status (in brackets proportion positive), 15/07/2021- 14/04/2022, Belgium.

| VOC | AG | Sex | Unvaccinated | Primary | Booster |
| --- | --- | --- | --- | --- | --- |
| Delta | 18-64 | F | 61,842 (0.38) | 269,224 (0.19) | 4,448 (0.12) |
|  |  | M | 52,934 (0.39) | 206,952 (0.20) | 2,476 (0.15) |
|  | 65-79 | F | 2,094 (0.46) | 30,006 (0.26) | 3,194 (0.13) |
|  |  | M | 1,359 (0.50) | 23,490 (0.32) | 2,954 (0.16) |
|  | >=80 | F | 588 (0.40) | 7,292 (0.23) | 3,532 (0.11) |
|  |  | M | 237 (0.40) | 4,572 (0.30) | 2,207 (0.17) |
| Omicron | 18-64 | F | 60,383 (0.64) | 119,972 (0.57) | 181,567 (0.40) |
|  |  | M | 51,153 (0.64) | 99,596 (0.57) | 130,586 (0.38) |
|  | 65-79 | F | 2,505 (0.65) | 3,162 (0.53) | 33,269 (0.42) |
|  |  | M | 1,511 (0.68) | 1,777 (0.56) | 27,034 (0.45) |
|  | >=80 | F | 668 (0.59) | 824 (0.46) | 18,014 (0.42) |
|  |  | M | 357 (0.63) | 411 (0.51) | 11,937 (0.50) |

**Table 2:** Numbers included (symptomatic infections) in the proportional hazard model by Variant of Concern (VOC), Age Group (AG), sex and vaccination status (in brackets proportion reported in the clinical hospital survey), 15/07/2021-14/04/2022, Belgium.

| VOC | AG | Sex | Unvaccinated |  | Primary |  | Booster |  |
| --- | --- | --- | --- | --- | --- | --- | --- | --- |
| Delta | 18-64 | M | 23,419 | (0.050) | 45,447 | (0.012) | 425 | (0.146) |
|  |  | F | 26,659 | (0.032) | 56,333 | (0.007) | 652 | (0.069) |
|  | 65-79 | M | 899 | (0.337) | 8,260 | (0.116) | 625 | (0.243) |
|  |  | F | 1,194 | (0.248) | 8,408 | (0.063) | 504 | (0.163) |
|  | >=80 | M | 186 | (0.586) | 1,866 | (0.311) | 474 | (0.281) |
|  |  | F | 383 | (0.475) | 2,057 | (0.213) | 465 | (0.206) |
| Omicron | 18-64 | M | 34,676 | (0.006) | 63,052 | (0.003) | 55,955 | (0.005) |
|  |  | F | 41,141 | (0.005) | 77,187 | (0.002) | 84,032 | (0.003) |
|  | 65-79 | M | 1,192 | (0.157) | 1,131 | (0.103) | 13,608 | (0.053) |
|  |  | F | 1,786 | (0.077) | 1,837 | (0.051) | 15,455 | (0.029) |
|  | >=80 | M | 353 | (0.425) | 281 | (0.285) | 7,269 | (0.163) |
|  |  | F | 558 | (0.323) | 479 | (0.221) | 8,914 | (0.111) |

### 4.2. VACCINE EFFECTIVENESS

Omicron was associated with reduced VEi and VEh-estimates compared to Delta. Initial Delta-VEi (0- 50 days) was 81% (CI 80-82) and Omicron-VEi was 37% (CI 34-40) for primary-vaccination. The initial VEi conferred by booster-vaccination was 84% (95%CI 83-85) against Delta and 52% (CI 52-53) against Omicron.

VEi for primary-vaccination decreased by 25 (Delta-VEi) and by 19 (Omicron-VEi) percentage points over a 100-150 day period corresponding to a Delta-VEi of 56% (CI 55-57) and an Omicron-VEi of 18% (CI 17-20). Because of insufficient follow-up before 2022 for most booster-vaccinated persons, we could not accurately assess waning of booster-vaccination against Delta. We observed waning against Omicron: Omicron-VEi was estimated at 25% (CI 24-27), 100-150 days after booster-vaccination, corresponding to a decrease of 27 percentage points from the initial Omicron-booster VEi-estimate.

VEh was significantly higher than VEi. Initial Delta-VEh was 93% (CI 91-95) for primary-vaccination and 94% (CI 93-94) for booster-vaccination. Against Omicron, initial VEh for primary-vaccination could only be estimated with large uncertainty (59% (CI 37-81)). Since the groups at high-risk of hospitalization were booster-vaccinated first, only few hospitalizations were registered among persons with a primary-vaccination for Omicron. For booster-vaccination, we estimated initial Omicron-VEh at 87% (CI 85-89). We observed a decrease of Omicron-VEh to 66% (CI 63-70), 100-150 days after booster-vaccination (Figure 1 & 2).

**Figure 1:**
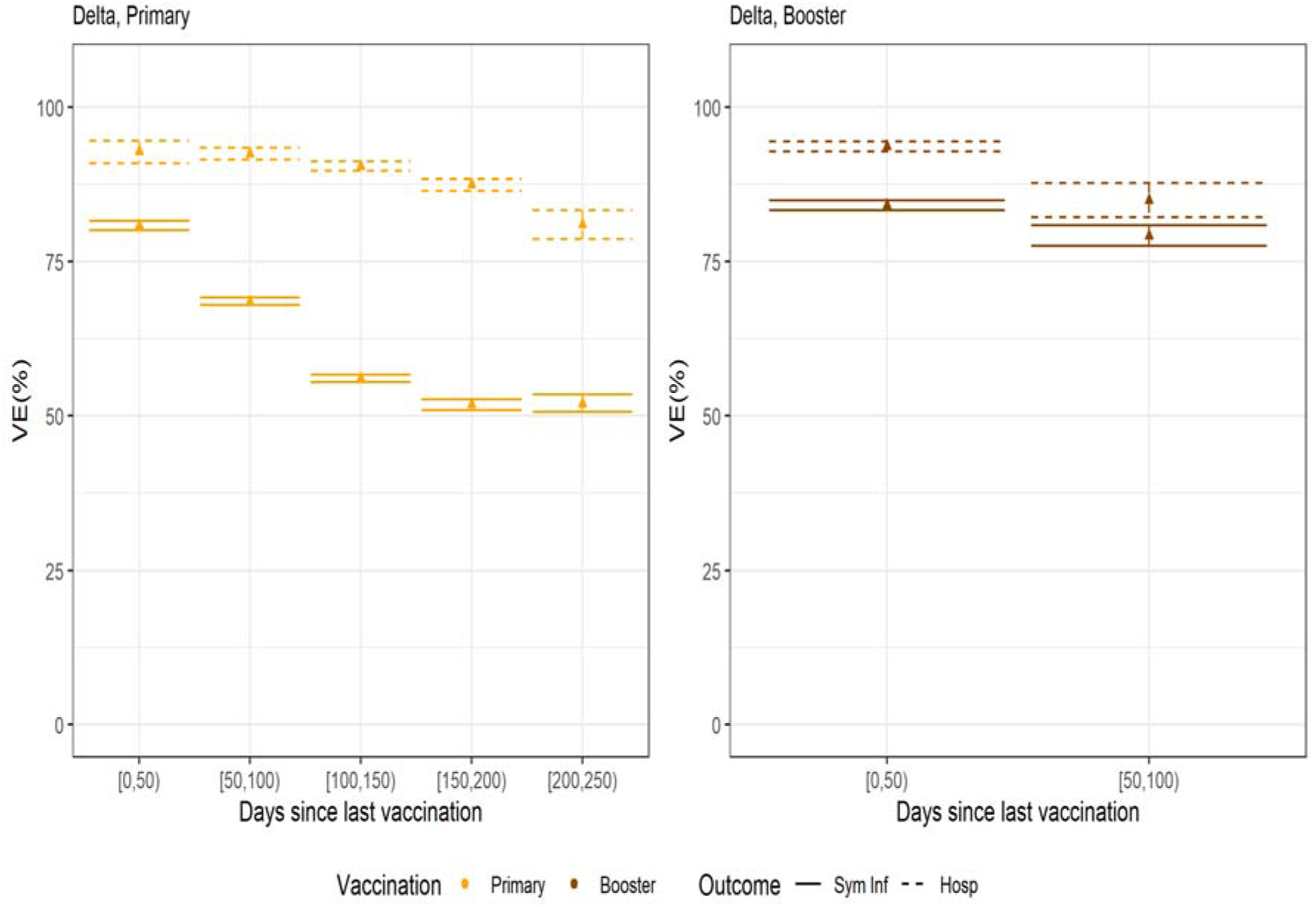
Vaccine Effectiveness against symptomatic infection (Sym Inf) and hospitalization (Hosp), adults, both sexes, (left) primary-vaccination, (right) booster-vaccination, 15/07/2022 – 06/12/2021 (period proxy for the Delta-VOC), Belgium.

**Figure 2:**
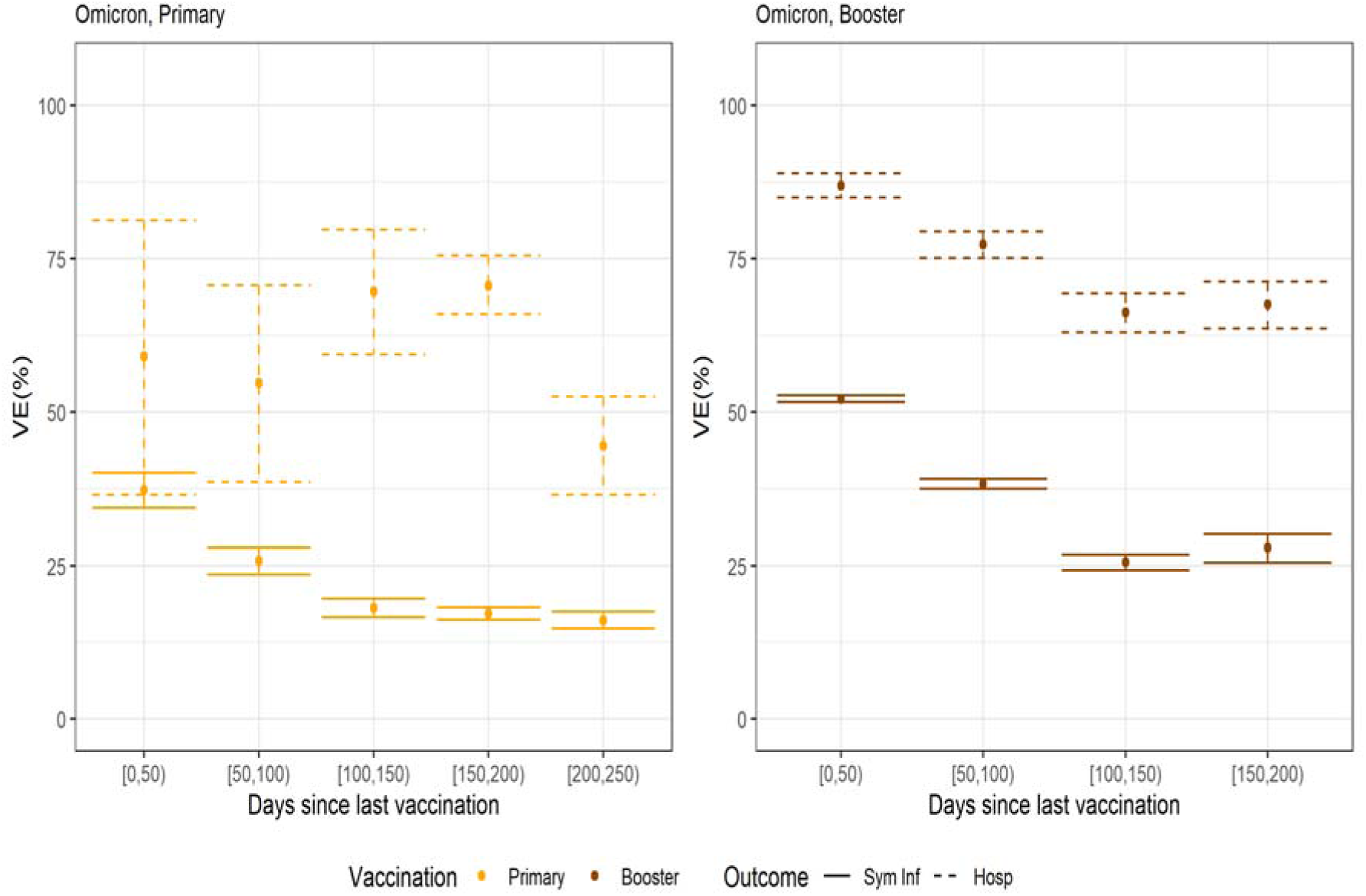
Vaccine Effectiveness against symptomatic infection (Sym Inf) and hospitalization (Hosp), adults, both sexes, (left) primary-vaccination, (right) booster-vaccination, 03/01/2022 – 14/04/2022 (period proxy for the Omicron-VOC), Belgium.

### 4.3. INFECTION-ACQUIRED AND HYBRID IMMUNITY AGAINST OMICRON

Protection against symptomatic Omicron-infections, conferred by a prior infection in unvaccinated persons, was higher for more recent infections. For prior infections from 2022, we estimated protection ((1-aOR)*100) at 76% (CI 72-80). For infections from the second half of 2021 at 40% (CI 38-48), for infections from the first half of 2021 at 21% (CI 19-24) and for infections from before 2021 at 14% (CI 11-16). Owing to the required 60-days interval between infections, prior infections from 2022 were only included for test results from March 2022 onwards.

Hybrid immunity also offered more protection if the last antigen exposure (either vaccination or infection) was more recent: VEi was estimated at 83% (CI 78-87) 0-50 days since booster-vaccination with a prior infection in 2022. While hybrid immunity waned at a rate comparable to primary-vaccination for prior infections from before the second half of 2021 and slower for more recent infections, booster-vaccination with prior infection continued to offer more protection than booster-vaccination without prior infection, even if the prior infection was from before 2021: 67% (CI 66-68) vs 52% (CI 52-53) 0-50 days after vaccination (Figure 3).

**Figure 3:**
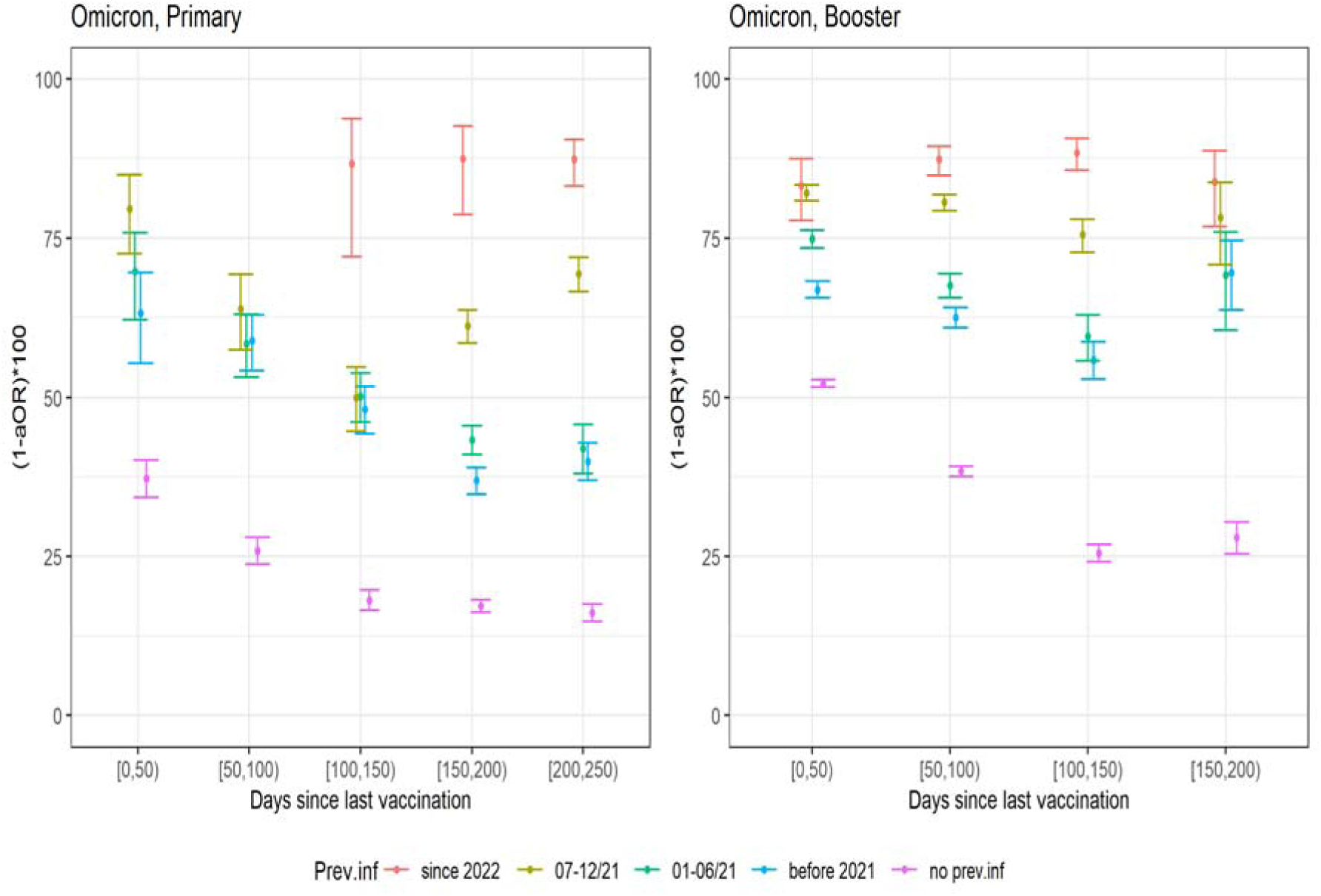
Hybrid immunity against symptomatic Omicron-infection (1- adjusted Odds Ratio)*100 by period of prior infection and days since vaccination for (left) primary-vaccination and (right) booster-vaccination, 03/01/2022 – 14/04/2022 (period proxy for the Omicron-VOC), Belgium.

Among hospitalized patients, 387 (2.8%) had a prior infection. For 171 persons, this was an infection from before 2021.

### 4.4. SEX AND AGE-DIFFERENCES

Overall CI of the VE-estimates by sex overlapped. Against Delta, in persons aged 65 years and more, 150-200 days after primary-vaccination, there is a significant difference in VEh by sex.. We observed significantly lower VEh in males 82% (CI 79-85) compared to females 88% (CI 86-90). No sex-specific statistically significant differences were found in younger age groups (<65 years), after booster-vaccination or for Omicron-infections (Figure 4).

**Figure 4:**
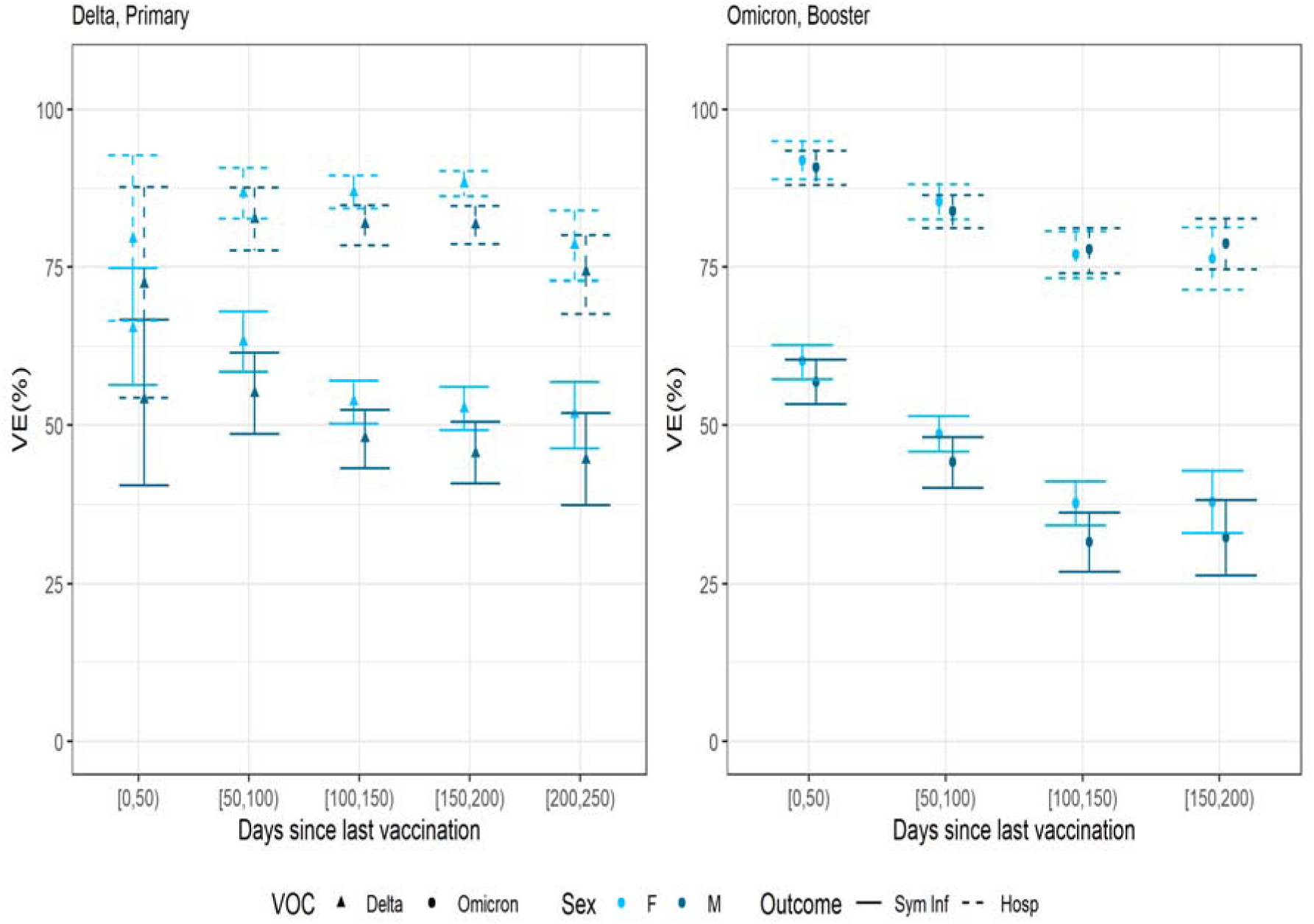
Vaccine Effectiveness against symptomatic infection (Sym Inf) and hospitalization (Hosp) for males (M) and females (F) in the age group 65 years and over, (left) primary-vaccination, Delta (right) booster-vaccination, Omicron, 15/07/2021-15/03/2022, Belgium.

Against Delta, VEi and VEh were higher in younger (below 65 years) compared to older (65 years and over) age groups. The difference was inverse (higher protection in older age groups) for booster-vaccination against Omicron (Figure 5). For both VOCs, the differences decreased as the time since vaccination increased.

**Figure 5:**
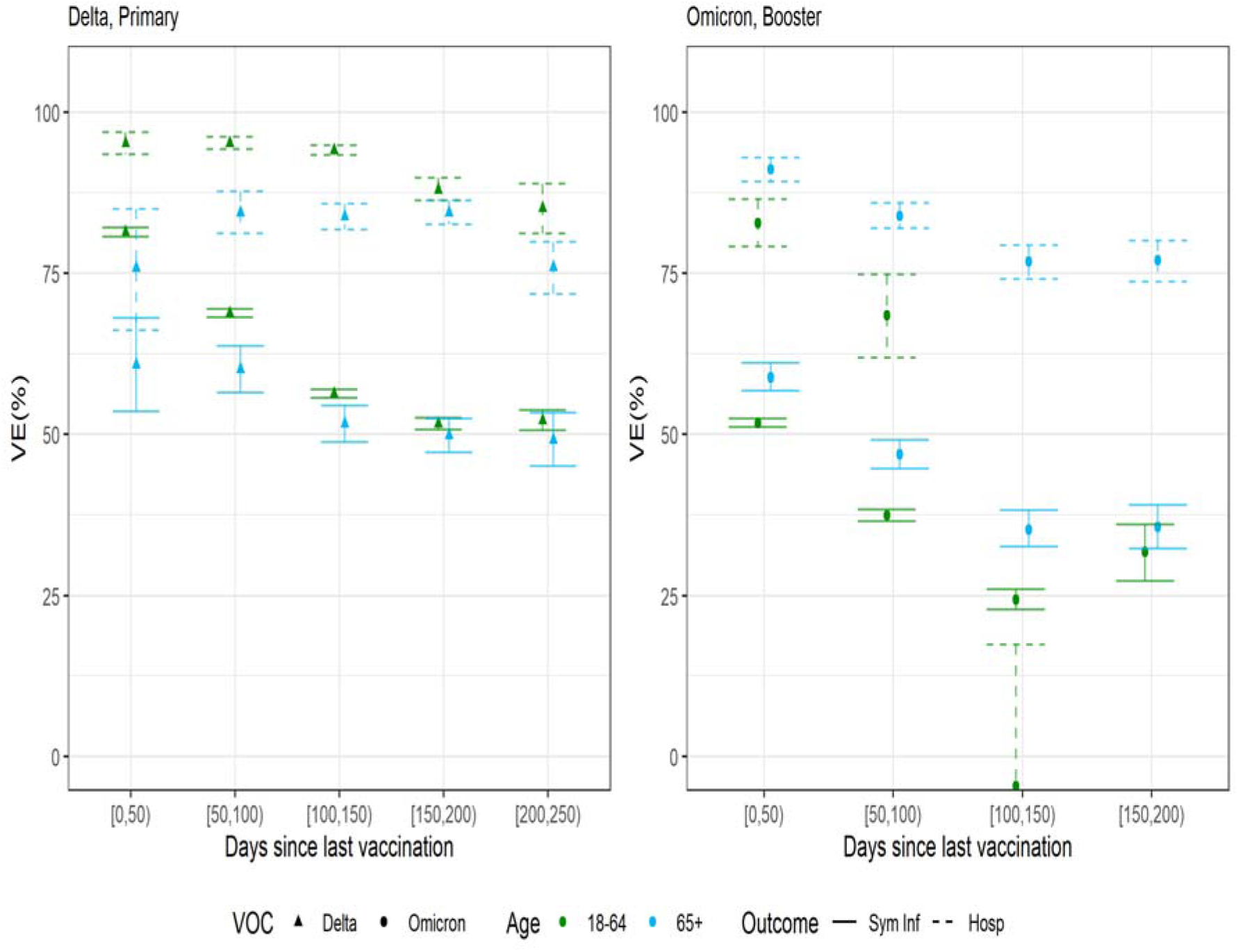
Vaccine Effectiveness against symptomatic infection (Sym Inf) and hospitalization (Hosp) for age groups below 65 years and 65 years and over, (left) primary-vaccination, Delta and (right) booster-vaccination, Omicron, 15/07/2021-14/04/2022, Belgium.

## 5. Discussion

### 5.1. SUMMARY

In this study, we presented the effect of vaccination and prior infection on the risk of SARS-CoV2 VOC-specific symptomatic infection and hospitalization. Our main findings include: (1) Omicron dominance and waning reduced VEi and VEh-estimates. (2) Booster-vaccination increased protection by restoring waning immunity and attenuating Omicron’s immune escape. (3) Hybrid immunity was associated with higher protection than vaccine-induced or infection-acquired immunity only. (4) While more recent infections and vaccinations were associated with higher protection against symptomatic infection, protection from the, within this study, oldest infections (over one year ago) and oldest vaccinations (200-250 days ago) remained significant.

### 5.2. OMICRON’S IMMUNE ESCAPE

Among the currently known VOCs, Omicron displays the most pronounced humoral immune escape [11,12]. We estimated the initial primary-vaccination VE against symptomatic Omicron-infections at 37% (CI 34-40). However, when Omicron became dominant at the start of 2022, Belgian adults without booster-vaccination had lower protection (VEi of 18% (CI 17-20)) because on average 120 days had passed since their primary-vaccination. Other studies have reported an initial Omicron-VEi of around 36% (preprint) [5,13] and no protection after >=180 days. Andrews et al. [6] reported initial Omicron-VEi of 49% (95%CI 47-50) declining to around 9% after 175 days for BNT162b2 primary-vaccination in the UK.

Shortly after the emergence of Omicron in South Africa, lower vaccine-induced protection against hospitalization was reported [14]. We estimated VEh at 70% (CI 59-80), significantly lower than the VEh-estimate against Delta of 90% (CI 90-91) (both 100-150 days after primary-vaccination). This might be linked to reduced reactivity in T-cell responses in some individuals [15]. From a public health perspective, it is important to state that, while the responses of the different immunity layers are VOC-specific [16], transmission and probability of severe outcomes are also VOC-specific. The public health impact of the lower VEh should be evaluated by the reported lower severity of Omicron compared to Delta in most age groups [17].

### 5.3. BOOSTER-EFFECTS

The waning of primary-vaccination Delta-VE-estimates reported in this study is in line with the 20-30% (VEi) and 9-10% (VEh) decreases reported for a six months period by a systematic review [18]. Booster-vaccinations restores waned protection and improved initial VEi-estimates against both VOCs. Our findings in older age groups are in accordance with *in-vitro* studies [19] and epidemiological studies which have estimated an initial Omicron-VEi of 60-75% after booster-vaccination (preprint) [5,20,21]. For younger age groups (<65 years) we report lower booster- vaccination VEi-estimates.

We estimated the initial Omicron-VEh around 87% (CI 85-89) after booster-vaccination. We observed waning of the VEh, after primary and booster-vaccination, albeit slower than waning of VEi. Other studies that included adequate follow-up time after booster-vaccination also made this observation [22–24]. USA-studies reported decreases from 91% to 78% and 85% to 55% by the fourth month after the third vaccine dose [25,26]. Towards the end of our follow-up time (200-250 days after the last vaccination dose) waning seemed to decelerate. The available serological evidence also describes stabilizing antibody titres six to nine months post-vaccination [27]. Future research should investigate the waning trend over longer time periods. This will not be straightforward as the high incidences associated with Omicron have impacted case-finding, e.g. in Belgium testing of high-risk exposure contacts was reduced from mid-January 2022 onwards, and undetected cases are a potential, typically downward, source of bias.

### 5.4. HYBRID IMMUNITY

In line with studies detecting antibodies [28,29] or significant VE-estimates [30] one year after infection, we observed low, but significant protection against symptomatic Omicron-infection by ‘over one year old’-infections in unvaccinated persons. In vaccinated persons, prior infections added to the vaccine-induced protection. The additional protection remained significant for the maximum follow-up included in this study. The UK SIREN-study also reported durable protection by old (up to 18 months) infections and subsequent vaccination [31]. The additional protection also remained after booster-vaccination. A quantitative benefit to neutralizing antibody responses has been observed after a fourth, heterologous, antigen exposure [27]. In contrast, repeated homologous antigen exposures might achieve maximal immunogenicity after three exposures [33,34]. We demonstrated that a more recent infection was associated with higher protection. This likely indicates both waning of infection-acquired immunity and differences in cross-neutralization between VOCs [35]. Given the functional relevance of immunological processes such as cross-neutralization and affinity maturation [11,36], observed time trends in convalescents are specific to the sequence of VOCs. Our results might therefore be most relevant to populations also confronted with an Alpha-Delta-Omicron sequence.

### 5.5. AGE & SEX

We have previously identified older males as the population with the lowest VE against Delta- infections, over time [1]. In this study, male sex was associated with a lower VEh-estimate against Delta for primary-vaccination, while no significant differences were visible after booster-vaccination. Our results with respect to age are less consistent. While, during the fourth wave (Delta-dominance), younger persons were associated with higher VE-estimates, the association inversed during the fifth wave (Omicron-dominance). Lower VE-estimates for younger age groups against Omicron have also been reported from the USA [26] and Denmark (preprint) [23]. While this observation can be linked to age-specific characteristics of Omicron or age-specific behavioral differences, a rise in undiagnosed infections during the fourth and fifth waves in younger populations, is another possible explanation.

## 5.6. LIMITATIONS

This study has several limitations.

With respect to the investigated outcomes; symptomatic infection is based on a self-reported date of symptom onset. There is only info on the occurrence of symptoms, without further information on the actual clinical presentation. We have tried to limit a bias from between-person heterogeneity with respect to self-reported presence of symptoms by including only one, randomly chosen, symptomatic period per person. With respect to hospitalization as outcome; no exhaustive patient registry is available for COVID-19-hospitalizations in Belgium. This study therefore is susceptible to sampling bias. In the present study, we assume no sampling bias by vaccination status. VEh-estimates should be compared with care between countries as countries’ hospital admission policies differ. While the included hospitalizations were for COVID-19 symptoms, we did not differentiate by severity or hospital outcome.

For prior infections, we did not make the distinction between symptomatic and asymptomatic presentation, while there is evidence that the protection differs by presentation [29]. We also did not differentiate between infections occurring before or after vaccination. Both have been reported to boost the quantity, quality and breadth of the humoral immune response [37,38]. We did differentiate prior infections by the time of their occurrence, but the periods we used were large and susceptible to confounding. For example, infections from the second half of 2021 were likely positively correlated with the time since vaccination. In addition, we can only include those prior infections that have been laboratory confirmed (either through PCR or (rapid) antigenic test). Given that case-ascertainment, and therefore detection of prior infections, might be age and sex-specific, comparison of VE’s between demographics should be done with care.

Differences in exposure by immunity status (because of higher circulation among contacts and/or behavior changes) remain a potential source of bias. To some extent the required use of a certificate (either vaccination, negative test or recovery), during the study period, to enter bars, restaurants, sport and cultural facilities might have caused additional exposure differences between vaccinated and unvaccinated persons. Future studies might opt for a reference category other than unvaccinated persons, but since this was the first Belgian study to estimate VEh, we chose a reference group (unvaccinated persons) that allowed for an intuitive interpretation. Using a test negative case-control design helped to control for differences in health seeking behaviour between vaccinated and unvaccinated persons. We did not include other variables that might affect exposure and testing behavior such as socio-economic status, profession, etc. We also did not include variables on individual susceptibility such as comorbidities or if a person was immunocompromised or not.

The sequential roll-out of the Belgian vaccination campaign as well as the booster campaign prioritizing the elderly has important implications for our study. Since we did not want to make any modelling assumptions (e.g. on the waning over time), the analyses are limited by the numbers available for each discrete time period. Relatively few younger persons received booster-vaccination before the Delta-period. Estimates of Delta-VE for booster-vaccination therefore are driven by effectiveness in older age groups and vice versa for primary-vaccination and Omicron. Consequences are (1) some estimates cannot be provided and the uncertainty around others is large, (2) estimates for all Belgian adults might be driven by certain age groups and not be representative for other age groups and (3) when comparing VE-estimates by time since vaccination for different age groups we are also comparing different calendar periods. We excluded minors from the analysis since they were not part of the targeted population for the initial vaccination campaign.

To limit the complexity of the study and increase its power, we did not differentiate between vaccine brands. Exploratory analysis were not pointing towards large differences between brands after booster-vaccination. For the primary schedule, we previously explored differences between brands[1].

In conclusion, we report significant immune-escape by Omicron for both symptomatic infection and hospitalization. Booster-vaccination attenuated the immune-escape and restored the loss in protection that resulted from waning. Hybrid immunity offered durable and high protection. While we observed waning of infection-acquired, vaccine induced and hybrid immunity, significant protection remained for the longest intervals (over one year) included in this study.

## Data Availability

Person-level data is not available, aggregated data from the present study can be obtained from the authors upon reasonable request.

https://epistat.wiv-isp.be/covid/

